# Key variants via Alzheimer’s Disease Sequencing Project whole genome sequence data

**DOI:** 10.1101/2023.08.28.23294631

**Authors:** Yanbing Wang, Chloé Sarnowski, Honghuang Lin, Achilleas N Pitsillides, Nancy L Heard-Costa, Seung Hoan Choi, Dongyu Wang, Joshua C Bis, Elizabeth E Blue, Alzheimer’s Disease Neuroimaging Initiative (ADNI), Eric Boerwinkle, Philip L De Jager, Myriam Fornage, Ellen M Wijsman, Sudha Seshadri, Josée Dupuis, Gina M Peloso, Anita L DeStefano, Alzheimer’s Disease Sequencing Project (ADSP)

**Author notes:** Corresponding authors Dr. Chloé Sarnowski, (editorial office correspondence) Dr. Anita L Destefano. Drs. Yanbing Wang and Chloé Sarnowski contributed equally to this work. Drs. Gina Peloso and Anita L DeStefano contributed equally to the supervision of this work.

## Abstract

**INTRODUCTION:** Genome-wide association studies (GWAS) have identified loci associated with Alzheimer’s disease (AD) but did not identify specific causal genes or variants within those loci. Analysis of whole genome sequence (WGS) data, which interrogates the entire genome and captures rare variations, may identify causal variants within GWAS loci.

**METHODS:** We performed single common variant association analysis and rare variant aggregate analyses in the pooled population (N cases=2,184, N controls=2,383) and targeted analyses in sub-populations using WGS data from the Alzheimer’s Disease Sequencing Project (ADSP). The analyses were restricted to variants within 100 kb of 83 previously identified GWAS lead variants.

**RESULTS:** Seventeen variants were significantly associated with AD within five genomic regions implicating the genes OARD1/NFYA/TREML1, JAZF1, FERMT2, and SLC24A4. KAT8 was implicated by both single variant and rare variant aggregate analyses.

**DISCUSSION:** This study demonstrates the utility of leveraging WGS to gain insights into AD loci identified via GWAS.

## 1. Introduction

Alzheimer’s disease (AD), the most common cause of dementia, has been ranked as the 6th leading cause of death in the United States and the 5th leading cause of death in older people (≥65 years old). Although the role of genetic factors in the development of AD has been widely recognized, genome-wide association studies (GWAS) typically identify regions or loci rather than specific genes and/or variants. Additionally, the loci identified by GWAS only explain a small portion of the total heritability of AD (h^2^ = [0.58–0.79]).^1^ Next-generation sequencing technology applied in diverse populations as part of the Alzheimer’s Disease Sequencing Project (ADSP) may help to elucidate the genetic architecture of AD, and thus, aid in the development of effective strategies to diagnose, prevent and treat AD.^2^

A recent large GWAS totalling 111,326 clinically diagnosed/’proxy’ AD cases and 677,663 controls has identified over 70 loci associated with AD and related dementias.^3^ However, the characterization of these loci remains incomplete. Leveraging whole genome sequence (WGS) data that encompasses the full spectrum of genetic variation including common and rare variants might identify important AD genes within these GWAS loci and provide a better understanding of the biological mechanisms involved in the pathophysiology of AD. Previous studies used WGS to identify genetic loci associated with AD.^3–6^ A family-based study conducted in 2,247 subjects from NIMH/NIA with replication in 1,669 independent participants from the ADNI/ADSP identified 13 novel AD candidate loci with rare-variant signals (FNBP1L, SEL1L, LINC00298, PRKCH, C15ORF41, C2CD3, KIF2A, APC, LHX9, NALCN, CTNNA2, SYTL3, and CLSTN2).^3^ More recently, the same team investigated association of groups of rare variants in the same datasets using a sliding-window approach and identified two novel genes (DTNB and DLG2) associated with AD.^4^ Additional studies conducted in Asian populations highlighted the importance of increasing representation of understudied population groups and value of WGS to uncover population-specific genetic loci.^5,6^

In this work, we focused on deep interrogation of known AD GWAS loci^7^ using the ADSP WGS data. The ADSP aims to identify protective or risk genetic contributors for AD in populations with diverse ancestry. The ADSP has generated single nucleotide variant and insertion/deletion (indel) calls based on WGS data from 4,789 participants, which are publicly available (R1 data release https://dss.niagads.org/datasets/ng00067-v1/). The goal of the current study is not replication of prior GWAS findings as we are underpowered to do so. In addition, the ADSP sample in the current analyses is not independent of the sample used in Bellenguez et al.^7^ Instead we aim to provide a more comprehensive look at GWAS loci.

We conducted single variant association analyses and rare variant aggregation association tests using the R1 WGS data of ADSP to identify specific genetic variants, genes and non-coding regions associated with AD within previously identified AD loci. We also examined multi-ancestry evidence for AD associations through population-specific analyses in White/European-ancestry (EA), Black/African-American (AA) and Hispanic/Latino (HI) subgroups, and a multi-population meta-analysis. The insights gained from our analysis will contribute to a better understanding of the AD pathogenesis and to potentially identify new targets for AD drug and treatment.

## 2. Methods

### 2.1 Study Participants

Data from the ADSP is available to qualified investigators via the National Institute on Aging Genetics of Alzheimer’s Disease Data Storage Site (NIAGADS) (https://dss.niagads.org/). This study was done under an approved NIAGADS research use statement and local Institutional Review Board approval. The current analyses focused on participants with WGS data in the NIAGADS file set named “R1 5K WGS Project Level VCF”. WGS data have been generated in multiple cohorts as part of the ADSP. The ADSP data included in this study are comprised of distinct phases including the Discovery, Discovery Extension, and Augmentation phases. The Discovery phase WGS was generated from individuals of multiplex AD families as previously described.^8–10^ The Discovery Extension phase consisted of a family component and a case control component. The Discovery Extension family component WGS was generated on additional members of selected families from the Discovery phase as well as members of 77 additional families. A set of 114 Hispanic control individuals was also sequenced with the family component.

A focus of the Discovery Extension case control component was to increase the diversity of the ADSP samples. The ADSP Discovery Extension WGS was generated on 3,082 individuals, with approximately one third from EA, AA, and HI populations. In the ADSP Discovery and Discovery Extension phases sequencing was performed at three sequencing centers via the National Human Genome Research Institute (NHGRI). Sequence data for ADSP Augmentation Studies were supported by NIA and private funding and are shared with the research community via NIAGADS. The ADSP data coordinating center, the Genomic Center for AD (GCAD), produced a jointly called and quality controlled (QC’ed) data set for WGS10 that included the ADSP WGS Discovery, Discovery Extension, and from the Augmentation phase, the Alzheimer’s Disease Neuroimaging Initiative (ADNI) study. Details of studies included in the ADSP can be found at NIAGADS under dataset: NG00067 ADSP Umbrella Study (https://dss.niagads.org/datasets/ng00067/).

### 2.2 WGS Quality Control

Low-quality variants were filtered out based on the GCAD provided flags, which were generated separately for the Family, Case-Control, and ADNI sub-studies.^10^ In addition, GCAD provided the ABHet ratio computed as (the total reference reads over all heterozygous genotypes)/(total alternative and reference reads over all heterozygous genotypes). A variant was excluded if it failed the GATK Variant Quality Score Recalibration (VQSR) filter, all genotypes were missing, was monomorphic, or if it had low call rate across all studies. Additional filtering was implemented within sub-study. If a variant had high read depth (>500 reads) within a study or had ABHet < 0.25 or ABHet > 0.75 within a sub-study, all the genotypes within that sub-study were set to missing. After these filters were applied, a final call rate filter of 95% across all sub-studies was implemented.

### 2.3 AD Phenotype Definition

The ADSP provides different AD status variable definitions for participants included via case-control versus family-based studies. In the current analysis, for individuals in the ADSP case-control study, we defined AD cases as individuals with either prevalent or incident AD. Case-control individuals with no prevalent or incident AD were defined as controls and those with missing status were defined as unknown. In the ADSP family phenotype file, possible values for the AD status variable include no dementia, definite AD, probable AD, possible AD, family-reported AD, other dementia, family reported no dementia, and unknown. For family-based individuals, we defined an AD case as either possible, probable or definite AD. AD controls were defined as individuals coded as no dementia. We redefined individuals with family-reported AD, other dementia, or unknown status as missing AD status. The ADNI phenotype data, which is part of the ADSP Augmentation study, provides information on mild cognitive impairment (MCI) in addition to AD status. Individuals with a current diagnosis of MCI (N=320) were included as AD controls in the current study. After selecting genetically unique individuals with AD status available, a total of 4,567 participants (2,383 controls and 2,184 cases) with WGS were included in the analyses.

### 2.4 Pooled sample single-variant association analysis

Single-variant association analysis of AD was performed on variants within GWAS loci for participants with both phenotype and genotype data available using GENESIS.^11^ Principal component analysis (PCA) was performed as described in the supplemental methods to assess and adjust for genetic ancestry of the study participants (**Figure A1**). The WGS samples included in the ADSP R1 WGS data set were sequenced across four sequencing centers (Baylor College of Medicine Human Genome Sequencing Center, The Broad Institute, McDonnell Genome Institute at Washington University School of Medicine, and Illumina) and 2 sequencing platforms (Illumina HiSeq 2000/2500, and Illumina HiSeq X Ten). In order to control for the effects from study design and technical differences, we generated indicator variables (study×sequencing center×sequencing platform) with 10 categories based on **Table 1**. We considered these indicator variables as technical covariates and defined case-control x Broad x HiSeq X Ten, which had the largest number of observations, as the reference group. We used a generalized logistic mixed-effects model to account for relatedness through a genetic relationship matrix (GRM). The GRM was estimated based on the same variants used in the PCA. We included sex, the technical covariates, and PC2 (based on a Bonferroni corrected significant p < 0.0016 for testing 32 PCs) as covariates in the null model. We performed the analysis across autosomes, and kept variants satisfying the criteria: call rate higher than 95% and minor allele count (MAC) higher than 20.

**Table 1.** Characteristics of the participants included in the ADSP R1 data set.

|  |  | **All (N=4,567)** | **AA\* (N=995)** | **HI\* (N=1,516)** | **EA\* (N=2,043)** | **Other (N=13)** |
| --- | --- | --- | --- | --- | --- | --- |
| **Age (sd)** |  | 76.9 (8.3) | 79.2 (7.6) | 75.1 (8.5) | 77.1 (8.4) | 77.3 (6.6) |
| **Alzheimer's Disease (%)** |  |  |  |  |  |  |
|  | **Case** | 2184 (47.8%) | 463 (46.5%) | 795 (52.4%) | 921 (45.1%) | 5 (46.2%) |
|  | **Control** | 2383 (52.2%) | 532 (53.5%) | 721(47.6%) | 1122 (54.9%) | 8 (53.8%) |
| **Sex (%)** |  |  |  |  |  |  |
|  | **Female** | 2822 (61.8%) | 710 (71.4%) | 1020 (67.3%) | 1086 (53.2%) | 6 (38.5%) |
|  | **Male** | 1745 (38.2%) | 285 (28.6%) | 496 (32.7%) | 957 (46.8%) | 7 (61.5%) |
| **Study (%)** |  |  |  |  |  |  |
|  | **ADNI** | 797 (17.5%) | 26 (2.6%) | 10 (0.7%) | 750 (36.7%) | 11 (84.6%) |
|  | **ADSP- case control** | 2963 (64.9%) | 944 (94.9%) | 1107 (73.0%) | 911 (44.6%) | 1 (7.7%) |
|  | **ADSP- family** | 807 (17.7%) | 25 (2.5%) | 399 (26.3%) | 382 (18.7%) | 1 (7.7%) |
| **Sequencing center (%)** |  |  |  |  |  |  |
|  | **Baylor** | 1241 (27.2%) | 0 (0) | 1103 (72.8%) | 138 (6.8%) | 0 (0) |
|  | **Broad** | 1263 (27.7%) | 2 (0.2%) | 286 (18.9%) | 974 (47.7%) | 1 (7.7%) |
|  | **Illumina** | 797 (17.5%) | 26 (2.6%) | 10 (0.7%) | 750 (36.7%) | 11 (84.6%) |
|  | **WashU** | 1266 (27.7%) | 967 (97.2%) | 117 (7.7%) | 181 (8.9%) | 1 (7.7%) |
| **Platform (%)** |  |  |  |  |  |  |
|  | **HiSeq X Ten** | 3227 (70.7%) | 965 (97.0%) | 1186 (78.2%) | 1074 (52.6%) | 2 (15.4%) |
|  | **HiSeq2000/2500** | 1340 (29.3%) | 30 (3.0%) | 330 (21.8%) | 969 (47.4%) | 11 (84.6%) |

\*AA: Black/African-American, HI: Hispanic/Latino; EA: White/European ancestry. Populations defined are described in the Methods.

To determine if significant variants identified provided distinct signals from the lead GWAS variants,^7^ conditional analyses were performed in loci displaying significant associations. Genotype data, coded as 0/1/2, for the lead GWAS variants in these loci was included in the null model in addition to the covariates. Association analyses conditioned on the lead GWAS variants were then rerun for the loci of interest.

### 2.5 Population specific association analysis

We conducted population specific analyses (null model and association analyses) for AD using GENESIS, accounting for genetic relatedness using a GRM. We defined three population groups (EA, AA, and HI). We selected a total of 2,144 EA participants based on PCA analysis performed using both the ADSP and the Human Genome Diversity Project (HGDP). Only participants who were not outliers based on 6 standard deviations (SD) from the mean for PCs 1-4 calculated in the European HGDP groups (Adygei, Basque, French, BergamoItalian, Orcadian, Russian, Sardinian, and Tuscan) were retained. We selected a total of 1,028 AA and 1,548 HI participants based on reported race and ethnicity. A total of 38 participants who identified as both African-American and Hispanic were placed in the Hispanic population. We included in the null model, in each population group, covariates associated with AD status at P ≤ 0.05. The EA null model included sex, ADSP family study status, Illumina sequencing center, HiSeq X Ten platform, PC 2, PC 9, and PC 15. The HI null model included sex, all sequencing centers, HiSeq X Ten platform, PC 13, PC 16, and PC 17. The AA null model included sex, Illumina sequencing center, and PC 1. We performed association analyses, in each population group, and retained the results with call rate higher than 95% and minor allele count (MAC) higher than 20. In addition, we performed a multi-population meta-analysis using three different models (fixed-effect, random-effect, and Han & Eskin’s modified random-effect) implemented in Metasoft^12^ by combining the population specific results satisfying the criteria of a within population MAC higher than 10. We then kept the meta-analysis results passing a total MAC across population groups higher than 20.

### 2.6 Gene-based tests

We tested the association of aggregate groups of low frequency (minor allele frequency (MAF) < 5%) or rare (MAF < 1%) genetic variants with AD status. Annotation for all called variants was generated using Ensembl VEP91 by the ADSP annotation working group. We selected missense or loss of function (lof) genetic variants based on the most severe variant consequence according to the ADSP Annotation WG Ranking Process and listed in the annotation file (frameshift variant, inframe deletion, inframe insertion, missense variant, protein-altering variant, splice acceptor variant, splice donor variant, start lost, stop gained, and stop lost). We conducted Sequence Kernel Association Test (SKAT, mmskat) and burden tests (combined multivariate and collapsing (CMC), emmaxCMC) with EPACTS (Efficient and Parallelizable Association Container Toolbox) using mixed-effect models adjusted for sex, technical covariates, and PCs significantly associated with AD status (PC2) accounting for genetic relatedness (GRM).

### 2.7 Non-coding rare variant analysis

For non-coding rare variant analysis, we used annotations from WGSA v0.8^13^ including annotations from ANNOVAR, VEP, SnpEff, COSMIC and SPIDEX. We conducted rare variant analysis using the variant-Set Test for Association using Annotation infoRmation (STAAR) method,^14^ which was developed to boost power of rare variant analyses by effectively incorporating both variant functional categories and multiple complementary functional annotations while accounting for relatedness and population structures. We used the same covariates (sex, technical covariates, and significant PCs) in the model as in single-variant analysis. The GRM was incorporated to account for relatedness among samples.

We aggregated sites that overlap enhancers and promoters around gene transcription start sites (TSS). The promoters within 5KB of a TSS that overlap DNase hypersensitivity sites (DHS) are defined as at least one WGSA H3K4me3 annotation for brain tissues (E067, E068, E069, E070, E071, E072, E073, E074, E081, E082), and the enhancers within 20KB of a TSS are defined by EnhancerFinder in Brain. We incorporated annotations from WGSA in the analysis, which include MAF, functional scores (GERP, GenoCanyon, RegulomeDB, FUNSEQ, CADD, Fathmm, EIGEN-PC), and the ENCODE score (DNASE). We then transformed the annotation scores to phred-scaled scores using -10×log10(rank(-score)/M), where M is the total number of variants tested in the analysis.

### 2.8 Focus on GWAS loci

Given the limited power to detect novel loci with the current sample size, we focused on exploiting WGS to provide insights on previously reported AD GWAS loci. We used the variants listed in Supplemental Table 5 in Bellenguez et. al as the previously reported AD GWAS top variants. For single variant association analyses, we looked up these lead variants in the ADSP WGS data. We then assessed ADSP WGS associations within 100KB of each lead GWAS variant. For gene-based and non-coding rare variant analysis, we obtained the results for genes or regions in the 100KB window around each lead variant. We included genes or non-coding regions for which any portion overlapped with the specified window. Using this paradigm, we identified 303 genes within 100kb of the index SNPs.

In general, we defined a threshold for statistical significance equal to 0.05/number of statistical tests and a suggestive threshold as 1/number of statistical tests. Within a 100kb window, many single variant tests were highly correlated. Therefore, we computed the effective number of independent tests using the simpleM approach^15^ and used the effective number of tests in the denominator when computing a window-specific threshold for single variant association testing. Effective number of tests were computed across the pooled sample and within each population sub-group (**Table A1**). **Figure 1** provides an overview of our analysis workflow.

**Figure 1.**
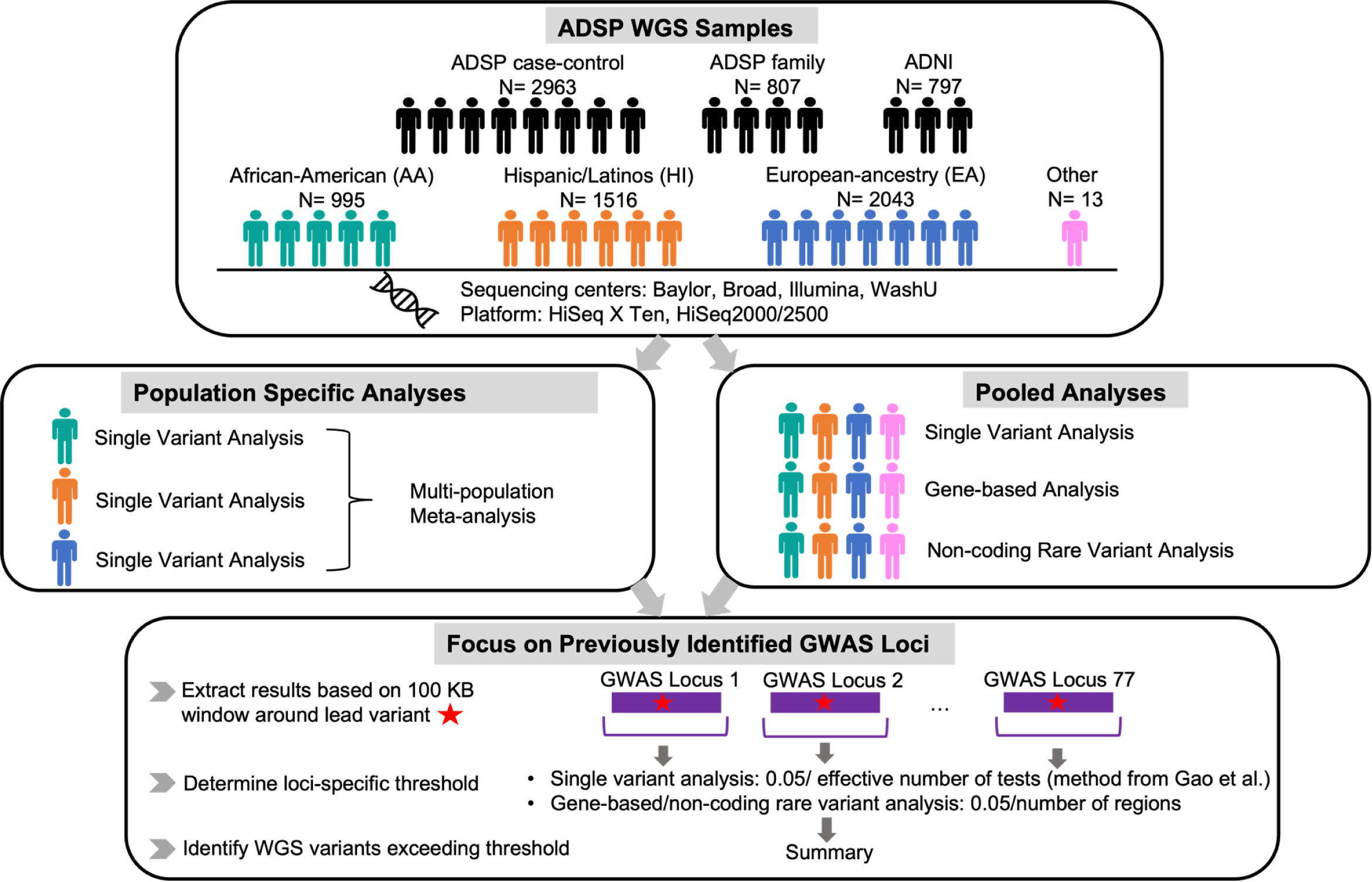
Schematic of the ADSP 5K analysis

We leveraged publicly-available multi-omic resource generated by applying quantitative trait locus (xQTL) analyses to RNA sequence and DNA methylation from the dorsolateral prefrontal cortex of 411 older adults from the Religious Orders Study (ROS) and Memory and Aging Project (MAP) studies^16^ to look-up the main genetic variants from the pooled association analysis.

## 3. Results

### 3.1 Description of ADSP data

After the QC of the ADSP data release NG00067.v2, there were over 95 million variants across 4,733 participants. A total of 4,567 individuals (2,383 controls, 2,184 cases) have available AD status and contributed to the analyses, among which 807 are from the ADSP family study, 2,963 are from the ADSP case-control study, and 797 are from ADNI. The participants included in the analyses were more likely to be women (61.8%) than men. The distribution of study design membership, sequencing centers, and sequencing platforms is summarized in **Table 1**.

### 3.2 Pooled sample single-variant association analysis

Genome-wide, there were about 20 million variants with call rate higher than 95% and MAC higher than 20 in the pooled sample analysis. Our model that included GRM and PC adjustments showed acceptable type-I error control (λ = 1.05, **Figure A2**). As expected, the strongest association was observed at the *APOE* locus, where the major *APOE* variant rs429358 (p = 7.2 × 10^-77^) was the top hit.

Among the specific lead GWAS variants from Bellenguez et al,^7^ none reached the strict significance threshold (p < 6 × 10^-4^, Bonferroni correction for the total number of variants tested) in the pooled sample association analysis. Using the suggestive significance threshold (p < 0.012; 1/83), we found associations for rs7401792 (p = 7.3 × 10^-4^, MAF= 49.2%) in the *SLC24A4* locus, rs75932628 (p = 3 × 10^-3^, MAF= 0.35%) in the *TREM2* locus, rs616338 (p= 4.4 × 10-3, MAF= 0.71%) in the *ABI3* locus, rs1358782 (p = 4.9 × 10^-3^, MAF= 22.9%) in the *RBCK1* locus, rs1160871 (p = 6.9 × 10^-3^, MAF= 40.2%) in the *JAZF1* locus, and rs602602 (p = 0.12, MAF= 27.7%) in the *MINDY2* locus. Full results for the 83 lead GWAS variants are provided in the supplement (**Table A2**).

Applying the significance thresholds based on the effective number of tests within 100 kb windows around the lead GWAS variants (**Table A1**), we identified 17 significant variants in the single variant association analysis in the pooled sample (**Table 2**). These 17 variants occur in five genomic regions on chromosomes 6, 7, 14, and 16. Forest plots for the top variant in each of these five regions are presented in **Figure 2**. Examination of linkage disequilibrium (LD) patterns show near perfect LD among the variants identified on chromosome 6, on chromosome 7 or for the one region on chromosome 14 with multiple variants. Only a single variant was identified on chromosome 16 and in one region on chromosome 14. Detailed LD information is provided in the supplement (**Figure A3**).

**Figure 2.**
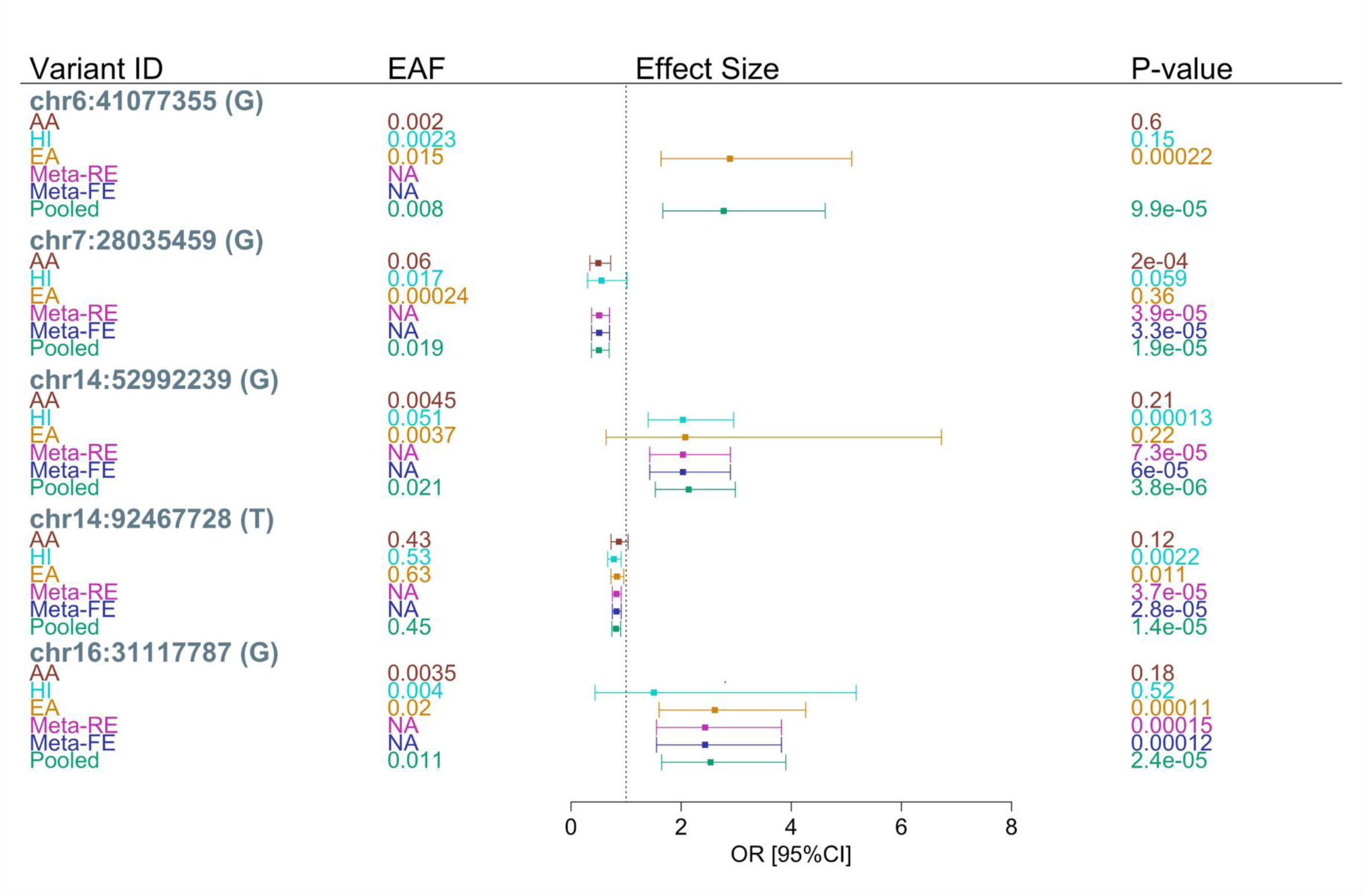
Top variants identified from single variant association analysis in the pooled sample within 100kb of the 83 lead GWAS variants Variant ID is in the form of chromosome:position (effect allele). Positions provided are on build 38. EAF is the effect allele frequency, Meta-RE is the multi-population meta-analysis using a random effect model, Meta-FE is the multi-population meta-analysis using a fixed effect model. The p-value for Meta-RE is calculated using Han and Eskin’s random effects model. The effect size and its 95% CI is not shown for variants with a minor allele count (MAC) < 10 in population specific analysis.

**Table 2.**
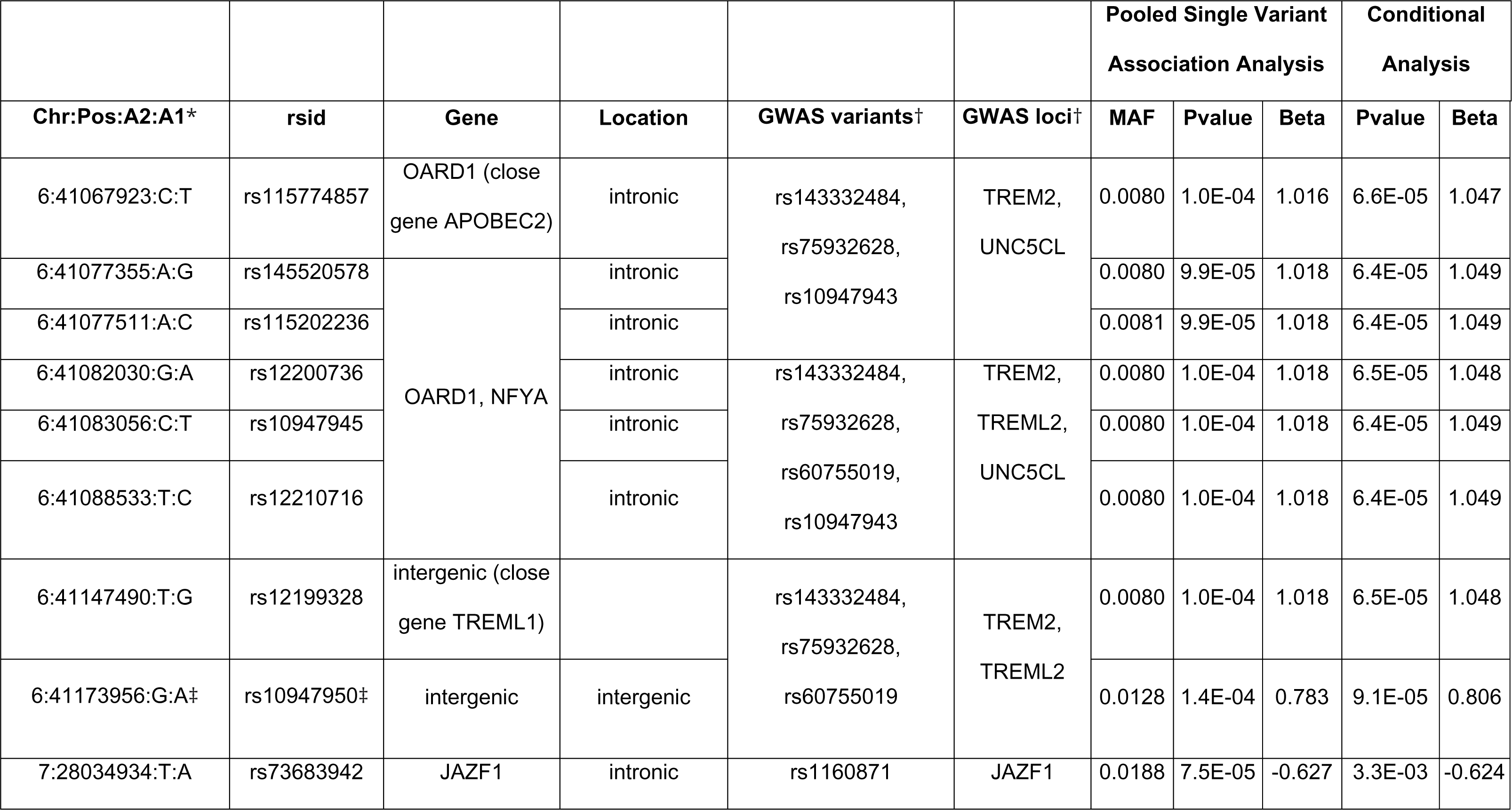
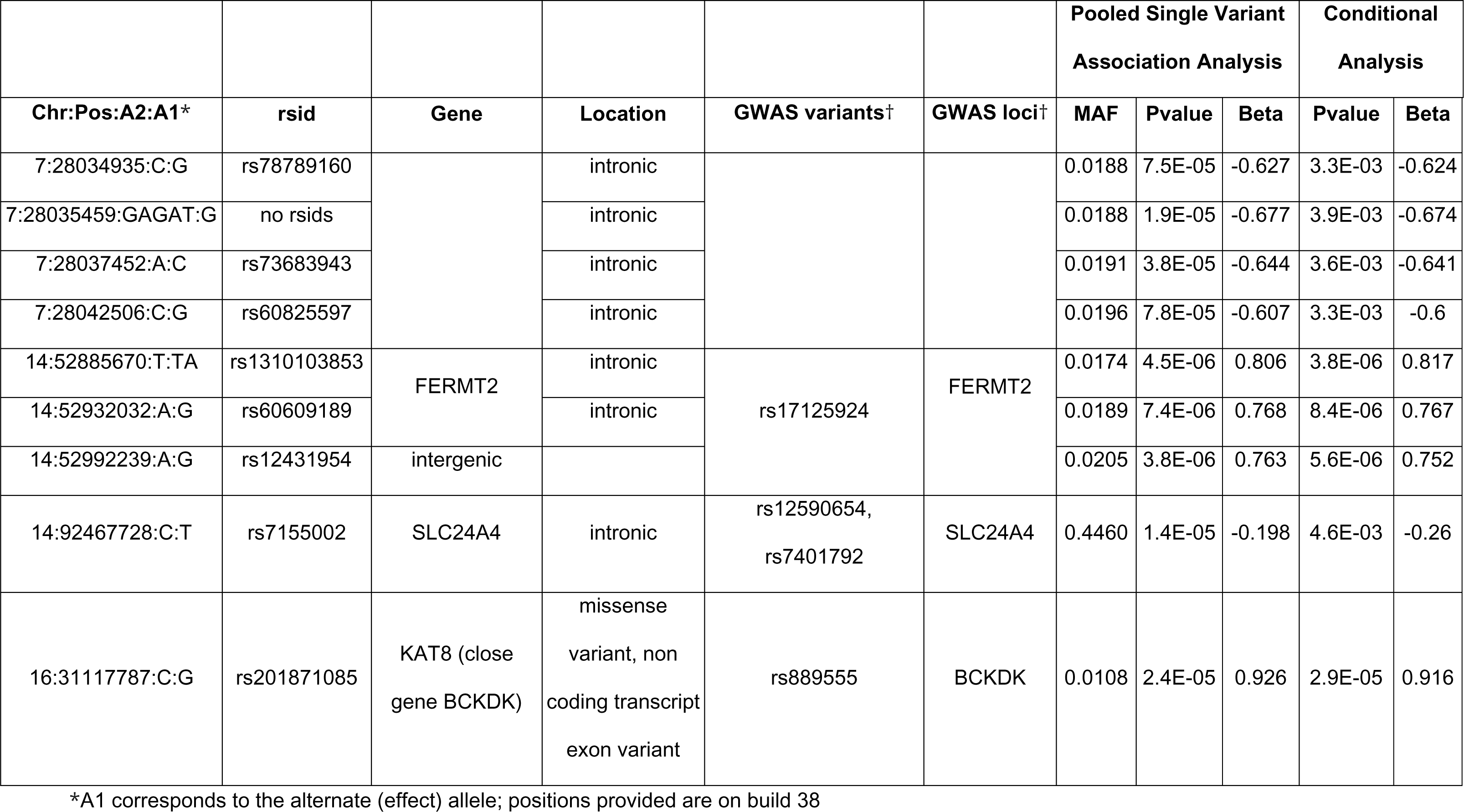
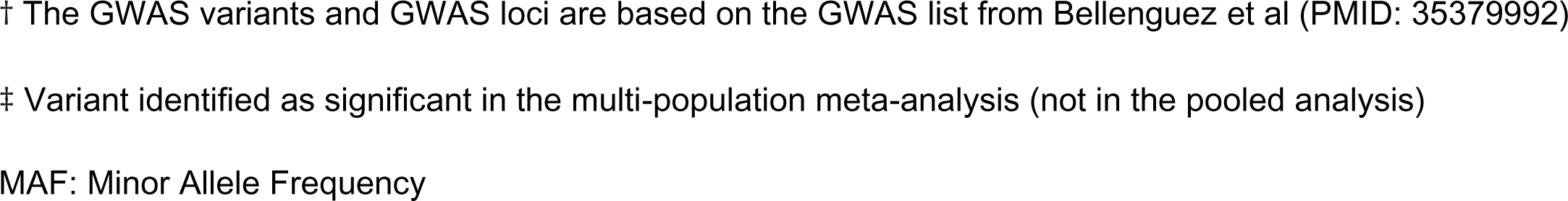
Significant variants from single variant association analysis in the pooled ADSP sample within 100kb of the 83 lead GWAS variants.

| Chr:Pos:A2:A1* | rsid | Gene | Location | GWAS variants† | GWAS loci† | Pooled Single Variant Association Analysis |  |  | Conditional Analysis |  |
| --- | --- | --- | --- | --- | --- | --- | --- | --- | --- | --- |
|  |  |  |  |  |  | MAF | Pvalue | Beta | Pvalue | Beta |
| 6:41067923:C:T | rs115774857 | OARD1 (close gene APOBEC2) | intronic | rs143332484,<br>rs75932628,<br>rs10947943 | TREM2,<br>UNC5CL | 0.0080 | 1.0E-04 | 1.016 | 6.6E-05 | 1.047 |
| 6:41077355:A:G | rs145520578 | OARD1, NFYA | intronic |  |  | 0.0080 | 9.9E-05 | 1.018 | 6.4E-05 | 1.049 |
| 6:41077511:A:C | rs115202236 |  | intronic |  |  | 0.0081 | 9.9E-05 | 1.018 | 6.4E-05 | 1.049 |
| 6:41082030:G:A | rs12200736 |  | intronic | rs143332484,<br>rs75932628,<br>rs60755019,<br>rs10947943 | TREM2,<br>TREML2,<br>UNC5CL | 0.0080 | 1.0E-04 | 1.018 | 6.5E-05 | 1.048 |
| 6:41083056:C:T | rs10947945 |  | intronic |  |  | 0.0080 | 1.0E-04 | 1.018 | 6.4E-05 | 1.049 |
| 6:41088533:T:C | rs12210716 |  | intronic |  |  | 0.0080 | 1.0E-04 | 1.018 | 6.4E-05 | 1.049 |
| 6:41147490:T:G | rs12199328 | intergenic (close gene TREML1) |  | rs143332484,<br>rs75932628,<br>rs60755019 | TREM2,<br>TREML2 | 0.0080 | 1.0E-04 | 1.018 | 6.5E-05 | 1.048 |
| 6:41173956:G:A‡ | rs10947950‡ | intergenic | intergenic |  |  | 0.0128 | 1.4E-04 | 0.783 | 9.1E-05 | 0.806 |
| 7:28034934:T:A | rs73683942 | JAZF1 | intronic | rs1160871 | JAZF1 | 0.0188 | 7.5E-05 | -0.627 | 3.3E-03 | -0.624 |
|  |  |  |  |  |  | MAF | Pvalue | Beta | Pvalue | Beta |
| 7:28034935:C:G | rs78789160 |  | intronic |  |  | 0.0188 | 7.5E-05 | -0.627 | 3.3E-03 | -0.624 |
| 7:28035459:GAGAT:G | no rsids |  | intronic |  |  | 0.0188 | 1.9E-05 | -0.677 | 3.9E-03 | -0.674 |
| 7:28037452:A:C | rs73683943 |  | intronic |  |  | 0.0191 | 3.8E-05 | -0.644 | 3.6E-03 | -0.641 |
| 7:28042506:C:G | rs60825597 |  | intronic |  |  | 0.0196 | 7.8E-05 | -0.607 | 3.3E-03 | -0.6 |
| 14:52885670:T:TA | rs1310103853 | FERMT2 | intronic | rs17125924 | FERMT2 | 0.0174 | 4.5E-06 | 0.806 | 3.8E-06 | 0.817 |
| 14:52932032:A:G | rs60609189 |  | intronic |  |  | 0.0189 | 7.4E-06 | 0.768 | 8.4E-06 | 0.767 |
| 14:52992239:A:G | rs12431954 | intergenic |  |  |  | 0.0205 | 3.8E-06 | 0.763 | 5.6E-06 | 0.752 |
| 14:92467728:C:T | rs7155002 | SLC24A4 | intronic | rs12590654,<br>rs7401792 | SLC24A4 | 0.4460 | 1.4E-05 | -0.198 | 4.6E-03 | -0.26 |
| 16:31117787:C:G | rs201871085 | KAT8 (close<br>gene BCKDK) | missense<br>variant, non<br>coding transcript<br>exon variant | rs889555 | BCKDK | 0.0108 | 2.4E-05 | 0.926 | 2.9E-05 | 0.916 |

\*A1 corresponds to the alternate (effect) allele; positions provided are on build 38
† The GWAS variants and GWAS loci are based on the GWAS list from Bellenguez et al (PMID: 35379992)
‡ Variant identified as significant in the multi-population meta-analysis (not in the pooled analysis)
MAF: Minor Allele Frequency

Conditional analyses were performed to determine if these associations represented the same signal as the lead GWAS variant from Bellenguez et al or a distinct signal. As shown in **Table 2**, the inclusion of the lead GWAS variant in the association model did not mitigate the association indicating the variants identified represent a distinct signal from the lead GWAS variants.

### 3.3 Population specific single-variant association analysis and multi-population meta-analysis

We conducted population specific association analyses in the three main subgroups (N=2,043 EA; N=995 AA, and N=1,516 HI participants). There was acceptable type-I error in the population specific analyses and the multi-population meta-analysis (**Figures A4-A7**). We confirmed the significant association of the *APOE* locus (rs429358) in both the population specific analyses and the multi-population meta-analysis. However, as found in previous studies^17,18^ the association was weaker in the Hispanic population (beta = 1.17 in EA, 1.02 in AA, and 0.59 in HI).

Population specific single variant analyses identified 23 significant variants in 11 loci within 100 kb of the lead GWAS variants (**Table 3**). Of these, 15 variants (8 loci) were identified in the EA population and 8 variants (3 loci) in the AA population. No significant variants were identified in the HI population. The only overlap in significant variants between the pooled sample and the population specific single-variant analyses was a missense variant in *KAT8*, which was very rare in the AA and HI subsamples (**Table A3**).

**Table 3.** Significant variants from single variant association analysis in the population subsamples within 100kb of the 83 lead GWAS variants.

|  | White / European Ancestry |  |  |  |  | Black / African American |  |  |  |  | Hispanic / Latino |  |  |  |  |
| --- | --- | --- | --- | --- | --- | --- | --- | --- | --- | --- | --- | --- | --- | --- | --- |
| Chr:Pos:A2:A1* | N | Freq | MAC | Pvalue | Beta | N | Freq | MAC | Pvalue | Beta | N | Freq | MAC | Pvalue | Beta |
| 4:11052797:T:C | 2040 | 0.0002 | 1 | 3.04E-01 | 2.069 | 984 | 0.137 | 270 | 8.37E-05 | -0.534 | 1513 | 0.059 | 180 | 4.94E-01 | 0.113 |
| 4:11053332:C:T | 2043 | 0.0002 | 1 | 3.04E-01 | 2.068 | 995 | 0.137 | 273 | 8.66E-05 | -0.530 | 1516 | 0.059 | 179 | 4.83E-01 | 0.116 |
| 5:180211637:C:T | 2042 | 0.143 | 584 | 7.14E-05 | 0.385 | 995 | 0.052 | 104 | 8.00E-02 | -0.351 | 1514 | 0.110 | 333 | 9.48E-01 | 0.009 |
| 5:180214978:G:A | 2042 | 0.143 | 586 | 5.37E-05 | 0.392 | 994 | 0.051 | 101 | 1.45E-01 | -0.296 | 1514 | 0.109 | 330 | 9.39E-01 | 0.010 |
| 5:180216117:C:T | 2041 | 0.143 | 582 | 3.69E-05 | 0.400 | 995 | 0.050 | 100 | 1.72E-01 | -0.278 | 1512 | 0.096 | 289 | 6.05E-01 | 0.073 |
| 5:180222862:G:A | 2042 | 0.144 | 590 | 3.70E-05 | 0.398 | 995 | 0.079 | 157 | 3.94E-01 | -0.144 | 1516 | 0.124 | 377 | 9.75E-01 | 0.004 |
| 5:180224704:C:T | 2041 | 0.145 | 591 | 2.85E-05 | 0.403 | 989 | 0.081 | 160 | 3.81E-01 | -0.147 | 1511 | 0.124 | 375 | 9.48E-01 | -0.008 |
| 8:11868930:C:G | 2043 | 0.007 | 27 | 6.52E-05 | 1.627 | 995 | 0.003 | 6 | 5.52E-01 | -0.494 | 1516 | 0.009 | 26 | 3.26E-01 | -0.414 |
| 11:86068255:T:G | 2042 | 0.810 | 774 | 6.19E-01 | 0.044 | 992 | 0.886 | 226 | 5.53E-05 | 0.576 | 1516 | 0.811 | 572 | 8.58E-01 | -0.019 |
| 11:86068268:A:G | 2042 | 0.810 | 774 | 6.19E-01 | 0.044 | 990 | 0.887 | 224 | 2.49E-05 | 0.607 | 1515 | 0.811 | 572 | 8.45E-01 | -0.021 |
| 11:86072833:A:G | 2042 | 0.815 | 755 | 6.67E-01 | 0.038 | 992 | 0.888 | 222 | 5.37E-05 | 0.577 | 1516 | 0.816 | 559 | 7.35E-01 | -0.037 |
| 11:86186529:G:A | 2038 | 0.142 | 577 | 8.10E-02 | 0.169 | 993 | 0.264 | 525 | 1.21E-04 | -0.403 | 1516 | 0.245 | 742 | 1.52E-01 | 0.137 |
| 14:105740487:C:T | 1863 | 0.524 | 1772 | 2.12E-05 | -0.479 | 981 | 0.308 | 605 | 5.33E-01 | 0.074 | 1446 | 0.430 | 1244 | 7.48E-02 | 0.204 |
| 15:63375962:G:A | 2042 | 0.0002 | 1 | 2.53E-01 | 2.318 | 994 | 0.101 | 200 | 1.18E-04 | 0.591 | 1514 | 0.035 | 105 | 9.71E-01 | 0.008 |
| Chr:Pos:A2:A1* | N | Freq | MAC | Pvalue | Beta | N | Freq | MAC | Pvalue | Beta | N | Freq | MAC | Pvalue | Beta |
| 15:63376200:A:G | 2042 | 0.0002 | 1 | 2.53E-01 | 2.317 | 994 | 0.106 | 210 | 1.24E-04 | 0.576 | 1515 | 0.040 | 122 | 7.96E-01 | -0.052 |
| 16:31117787:C:G | 1917 | 0.020 | 76 | 1.11E-04 | 0.961 | 991 | 0.004 | 7 | 1.77E-01 | 1.033 | 1489 | 0.004 | 12 | 5.16E-01 | 0.412 |
| 16:86357432:T:C | 2043 | 0.088 | 359 | 3.53E-05 | -0.497 | 995 | 0.132 | 262 | 7.55E-01 | -0.043 | 1516 | 0.097 | 293 | 4.62E-01 | 0.100 |
| 17:46724128:T:C | 2043 | 0.007 | 30 | 3.58E-05 | 1.607 | 995 | 0.002 | 3 | 5.80E-01 | -0.643 | 1515 | 0.006 | 17 | 3.75E-01 | 0.514 |
| 17:46747538:C:T | 2043 | 0.007 | 28 | 3.69E-05 | 1.649 | 994 | 0.002 | 3 | 5.79E-01 | -0.645 | 1515 | 0.006 | 17 | 3.76E-01 | 0.513 |
| 17:58269710:G:A | 2042 | 0.016 | 67 | 7.96E-05 | 1.069 | 995 | 0.004 | 8 | 7.23E-01 | 0.256 | 1515 | 0.005 | 14 | 7.84E-01 | -0.169 |
| 20:56407698:G:A | 2037 | 0.004 | 16 | 3.43E-05 | 2.328 | 991 | 0.001 | 2 | 8.15E-01 | 0.339 | 1516 | 0.002 | 6 | 8.86E-01 | -0.172 |
| 20:56490678:G:T | 2041 | 0.005 | 19 | 4.74E-05 | 2.021 | 995 | 0.002 | 4 | 4.93E-01 | -0.703 | 1516 | 0.002 | 6 | 8.86E-01 | -0.172 |
| 20:56505267:G:A | 2041 | 0.005 | 20 | 1.57E-05 | 2.095 | 995 | 0.002 | 4 | 4.93E-01 | -0.703 | 1513 | 0.002 | 6 | 8.83E-01 | -0.177 |

\*A1 corresponds to the alternate (effect) allele; positions provided are on build 38
Variants with a minor allele count (MAC) < 10 in a population subsample were not included in the meta-analysis

### 3.4 Gene-based tests

QQ plots for the SKAT and burden tests are provided in **Figure A8**. Using the multiple-testing correction threshold for 303 genes (p < 1.7 × 10^-4^), *KAT8* (p = 2.2 × 10^-5^, MAF< 5%) lying within 100 KB of a GWAS variant was detected to be significantly associated with AD status by SKAT, and it was also shown as a suggestive association (p < 3.4 × 10^-3^) using CMC (p = 9.2 × 10^-4^). SKAT detected suggestive associations in *LAIR1* (p = 0.0023, MAF < 5%) and *ATF5* (p = 5.7 × 10^-4^, MAF < 5%) within 100KB. CMC identified *TREM2* (p = 3.3 × 10^-3^ for MAF < 1% and 8.9 x 10^-4^ for MAF< 5%) within 100KB of GWAS variants.

### 3.5 Non-coding rare variant analysis

QQ plots showed deflated type-I error (λ = 0.75), most likely due to small sample size, in the STAAR rare-variant analysis. (**Figure A9**). No regions were identified as significant using the STAAR approach. The top STAAR results overlapping GWAS loci are shown in **Table A3**.

### 3.6 xQTL analysis lookup

We did not identify significant mQTL or eQTL associations for the main genetic variants identified in the pooled analysis. We could not look up some of the less frequent variants on chr7, 14 and 16 in the QTL results as these analyses were restricted to common variants. Suggestive associations have been reported between rs10947950 (chr6) and cg25473438 (beta = 0.24, P = 1.4 × 10^-8^) and between rs7155002 (chr14) and cg12072028 (beta = 0.19, P = 5.8 × 10^-6^), **Table A4**. The CpG cg12072028 is located in the intron 1 of *RIN3* and modest associations have been described between rs7155002 and *RIN3* expression in the brain (beta = 0.04, P = 0.01), **Table A4**.

## Discussion

GWAS have been essential in identifying genetic loci associated with AD. However, GWAS loci typically contain scores of genes and thousands of variants. Additional studies are needed to pinpoint specific genes or variants as the ones influencing risk for AD. WGS provides complete genomic sequence and hence enumerates both common and rare variants. WGS therefore has the potential to provide information beyond common variants that are the cornerstone of GWAS. In the current study, we have examined WGS from the ADSP R1 data set focusing on previously implicated regions to better understand important variants within AD GWAS loci in a diverse study sample. We identified 17 significant variants in five genomic regions using single variant association analysis in the pooled sample. The majority of these variants were intronic, although two intergenic and one missense variant were also identified. Bellenguez et al^7^ identified multiple lead GWAS variants on chromosome 6, which yielded overlapping 100kb windows defined by our approach. We identified seven significant variants on chromosome 6 that are in nearly complete LD. Six of these variants are located within intronic regions of *OARD1* and *NFYA* genes, and one variant is very close to *APOBEC2*. One variant was intergenic with the closest gene being *TREML1*. This region contains *TREM2*, for which several rare coding variants have been implicated as conferring risk for AD.^19–22^ *TREM2* showed suggestive evidence of association in gene-based analyses indicating multiple variants in this region are likely to play an important role in AD. The missense variant rs75932628 was one of the lead GWAS variants from Bellenguez et al. and has been identified as a functional variant for AD.^23–25^ The variant rs75932628 has an MAF=0.0035 in the ADSP pooled sample and p=0.003 for single variant association with AD. This suggestive association is driven by the EA population (MAF=0.007) as this variant is less frequently observed in the AA (MAF=0.001) or HI (MAF=0.0003) populations. Our conditional analysis indicates that the variants we identified implicating *OARD1/NFYA/TREML1* have a distinct effect from rs75932628. *OARD1* encodes a deacylase with a function to catalyse O-acetyl-ADP-ribose during multiple cellular processes. A homozygous mutation could lead to cell death and cause a form of childhood neurodegenerative disorder.^26^ *NFYA* encodes a subunit of nuclear transcription factor Y, which is a ubiquitous transcription factor. The gene is involved in post-transcriptional regulation with tissue-specific preference, and it is suppressed in the brain of model mice with Huntington’s disease^27^ and spinal and bulbar muscular atrophy.^28^ *TREML1* encodes a protein belonging to the family of triggering receptors expressed on myeloid cells-like (TREM). A deficiency of *TREML1* might result in haemorrhage due to localized inflammatory lesions.^29^

The five significant variants on chromosome 7 are in a strong LD block and all variants are intronic for *JAZF1*, which encodes a transcriptional repressor. The gene has been linked with diabetes mellitus and cancer, but also has a role in lipid metabolism supporting the connection between lipid levels and AD.^30^ The *JAZF1* GWAS variant (rs1160871) is a strong eQTL in microglia and considered a Tier 1 (highly plausible) AD gene.^7^ The significance of the variants at this locus was slightly attenuated by conditioning on the nearby GWAS variant (rs1160871), indicating that this may be a shared effect with the lead GWAS variant.

We identified two regions on chromosome 14, with significant variants intronic to *FERMT2* and *SLC24A4*. The association of the intronic variant rs7155002 for *SLC24A4* was slightly attenuated by the conditional analysis indicating that this may be a shared effect with the lead GWAS variant. Lookup in brain xQTL data shed light on potential biological regulatory mechanisms in *RIN3* that has also been implicated in AD.^31,32^ *FERMT2* encodes plekstrin homology domain-containing family C member 1 and is known to be involved in APP metabolism.^33^ The under-expression of *FERMT2* was associated with mature APP level increment in the cell surface.^33^ Previous studies reported that *FERMT2* is also involved in cardiac and skeletal muscle development^34^ and cancer progression.^35,36^ *SLC24A4* encodes a member of the potassium-dependent sodium/calcium exchanger protein family and is associated with neural development.^37^ A homozygous mutation in *SLC24A4* may cause amelogenesis imperfecta^38,39^ but the function of *SLC24A4* in AD is not clear yet.

A rare missense variant (rs201871085, MAF = 0.0108 in the pooled sample) within *KAT8* (lysine acetyltransferase 8) on chromosome 16 was significantly associated with AD. *KAT8* was also significant in the low-frequency variant gene-based analyses. *KAT8* encodes a member of the MYST histone acetylase protein family that has a characteristic MYST domain containing an acetyl-CoA-binding site, a chromodomain typical of proteins which bind histones, and a C2HC-type zinc finger. This gene has been recently identified by two large-scale GWAS of clinically diagnosed AD and family history of AD^40,41^ and by a novel knockoff method when applied to the ADSP Data.^42^ Aberrant expression patterns of *KAT8* might be associated with AD progression.^43^ *KAT8* appears like a promising candidate gene that is involved in cerebral development^44^ and may play a role in neurodegeneration in both AD and Parkinson’s disease.^45,46^ We were not able to look-up the variant rs201871085 in the brain xQTL data. This variant might not have been analyzed due to a low frequency or a low quality of imputation, thus highlighting the importance of leveraging whole genome sequence data. The ADSP represents a diverse population sample, although in this early release of ADSP WGS data the sample size within a specific population is limited (N_EA_ = 2,043, N_AA_ = 995, N_HI_ = 1,516). Population specific analyses provide information about patterns of allele frequency for AD associated variants among populations. Among the five loci identified as significant in the pooled single variant association analysis, two regions (chr6 & 16) displayed EA-specific associations and corresponded to low frequency variants in EA that were rare in other population groups. One signal (chr7) was driven by a variant common in AA with a low frequency in HI, and rare in EA. Finally, two regions (chr14) were driven by HI signals with one variant common in all population groups, and one variant common in HI but rare in EA and AA. All these results are summarized in **Table A3**.

The signals identified only in AA in the population specific analyses (chr4 and 14) corresponded to SNPs common in AA that were less common in HI and rare in EA. A few signals identified only in EA corresponded to variants that were rare in all population groups (chr8, 17 and 20). Two low frequency signals identified only in EA (chr16 and 17) corresponded to SNPs that were rare in AA and HI. Finally, three signals identified only in EA (chr5, 14, and 16) corresponded to SNPs that were common in different population groups. All these results are summarized in **Table 3**.

A strength of this study is the analysis of WGS data that was jointly called and QC’ed by a single data coordinating center. The diversity in genetic ancestry of the participants included is another strength. Despite this diversity, a limitation of the study is the moderate sample size within each population analysed. To overcome this limitation, main analyses were focused on the pooled sample. The ADSP is ongoing with larger WGS data sets being publicly released and planned. Future analyses with larger sample size may yield additional insights, specifically for population specific effects. The current study demonstrates the importance of leveraging whole genome sequence data to gain insights into loci identified via GWAS and highlights the contribution of low frequency variants to AD risk.

## Data Availability

Data from the Alzheimer's Disease Sequencing Project (ADSP) is available to qualified investigators via the National Institute on Aging Genetics of Alzheimer's Disease Data Storage Site (NIAGADS) (https://dss.niagads.org/).

## Abbreviations

(AD): Alzheimer’s disease
(ADSP): the Alzheimer’s Disease Sequencing Project
(AA): Black/African-American
(CMC): Combined Multivariate and Collapsing
(EA): White/European-ancestry
(GWAS): Genome-Wide Association Studies
(GRM): Genetic Relationship Matrix
(HI): Hispanic/Latino
(LD): Linkage Disequilibrium
(MAC): Minor Allele Count
(MAF): Minor Allele Frequency
(MCI): Mild Cognitive Impairment
(PCA): Principal Component Analysis
(QC): Quality Control
(SKAT): Sequence Kernel Association Test
(STAAR): variant-Set Test for Association using Annotation infoRmation
(WGS): Whole Genome Sequencing

## Acknowledgments

We thank the contributors who collected samples used in this study, as well as patients and their families, whose help and participation made this work possible.

<u>ADSP</u>: Data for this study were prepared, archived, and distributed by the National Institute on Aging Alzheimer’s Disease Data Storage Site (NIAGADS) at the University of Pennsylvania (U24-AG041689), funded by the National Institute on Aging (accession NG00067). The full acknowledgement statement for the ADSP can be found at: https://dss.niagads.org/datasets/ng00067/

<u>ADNI</u>: Data used in preparation of this article were obtained through NIAGADS. The investigators within the ADNI contributed to the design and implementation of ADNI and/or provided data but did not participate in analysis or writing of this report. A complete listing of ADNI investigators can be found at: http://adni.loni.usc.edu/wp-content/uploads/how_to_apply/ADNI_Acknowledgement_List.pdf

