## Supplemental Text and Figures for "Key variants via Alzheimer’s Disease Sequencing Project whole genome sequence data"

### Supplementary Text

### Principal component analysis (PCA) methods

To visualize and understand the complex population structure we conducted principal component analysis (PCA) using the ADSP R1 whole genome sequence (WGS) data. Hardy-Weinberg Equilibrium (HWE) is a key filter for selecting variants for PCA. To have a more robust estimate of HWE for sequence-based genotypes under complex population structure, Robust Unified HWE Test (RUTH) was implemented.[1] RUTH requires PCs for implementation. Therefore, the first step was to perform PCA for RUTH. We used the Human Genome Diversity Project (HGDP) data as a reference data panel to generate principal components (PCs) that were later used for RUTH calculation. We first performed initial filtering of the HGDP data, restricting to variants with call rate > 95% and minor allele frequency (MAF) > 5%. We then merged the filtered HGDP data with the ADSP R1 WGS data and selected the overlapping variants across the two datasets. We finally restricted the variants to those who satisfy the criteria of call rate > 99%, MAF > 5%, and linkage disequilibrium (LD) with a threshold of squared correlation ($R^2$) < 0.1 in a sliding 50KB window for PCs calculation. The first 20 PCs were then used as the input in RUTH calculation. Moreover, we used unrelated samples as identified by GENESIS to calculate RUTH-HWE.

We then performed a second PCA using only the ADSP R1 WGS data to generate PCs for use in association analysis. We applied the following criteria to identify variants used in PCA: call rate > 99%, MAF > 0.05, LD with a threshold of $R^2 = 0.1$ in a sliding 50KB window, and RUTH-HWE p-value < $10^{-4}$. The RUTH-HWE was estimated as described above, and we used PLINK2 to filter the genetic data on call rate, MAF, and LD.[2] The PCA included 4,733 individuals who passed sample quality control (QC) and with available WGS data in the ADSP.

We used an iterative procedure combining PC-AiR and PC-Relate in GENESIS to calculate 32 PCs, as this approach is robust to the population structure and admixed samples.[3] We ran the iteration two times to simultaneously estimate PCs following the steps described elsewhere (https://uw-gac.github.io/topmed_workshop_2017/). A total of 213,539 variants were identified after filtering. Among 4,733 QC'ed samples included in the analysis, 3,310 unrelated samples were identified by the threshold of kinship coefficient < $2^{-11/2}$ (corresponding to $4^{th}$ degree relatives) and were used for PC calculation.

**Supplementary Tables are large and thus are provided in Excel documents**

**Supplementary Figures**

**Figure A1**. PC1 vs PC2 of all 4,733 whole genome sequence (WGS) samples, color-coded by ancestry/ethnicity (left), or by studies and platforms, and symbolled by sequencing centers (right)

**Figure A2**. QQ-plot of the single-variant association analysis across WGS in the ADSP

**Figure A3**. LD plots for each of the main GWAS regions with several signals detected with the pooled analysis. LD was calculated using the ADSP data in each population group separately.

**Figure A4**. QQ-plot of the single-variant association analysis performed in the White/European Ancestry group

**Figure A5**. QQ-plot of the single-variant association analysis performed in the Black/African American group

**Figure A6**. QQ-plot of the single-variant association analysis performed in the Hispanic/Latino group

**Figure A7.** QQ-plots for the multi-population meta-analysis under different models.

**Figure A8**. QQ plots for the rare-variant aggregation analyses (gene-based) conducted with EPACTS based on missense and loss of function rare genetic variants.

**Figure A9**. QQ plots for the non-coding rare-variant aggregation analysis using STAAR.

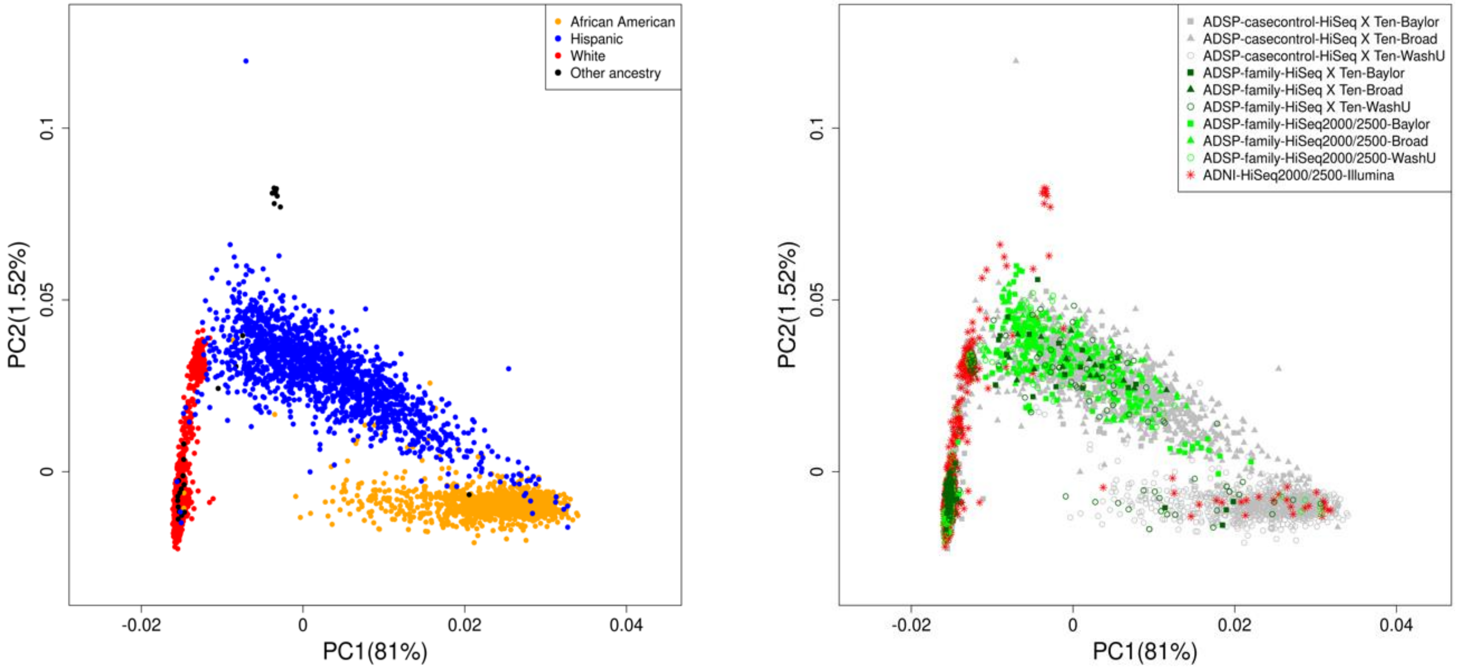


**Figure A1**. PC1 vs PC2 of all 4,733 whole genome sequence (WGS) samples, color-coded by ancestry/ethnicity (left), or by studies and platforms, and symbolled by sequencing centers (right)

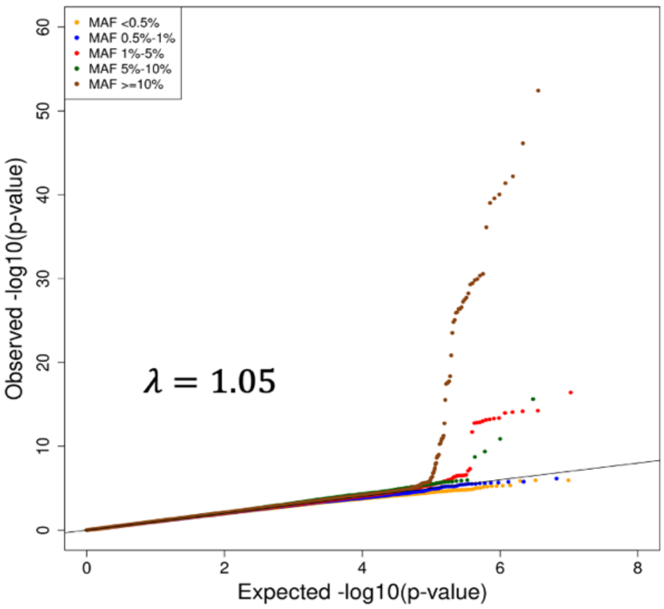


**Figure A2**. QQ-plot of the single-variant association analysis across WGS in the ADSP

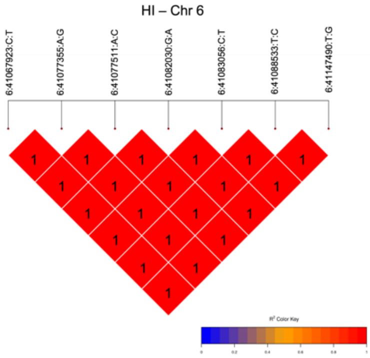
HI – Chr 6
6:41067923:C:T
6:41077355:A:G
6:41077511:A:C
6:41082030:G:A
6:41083056:C:T
6:41088533:T:C
6:41147490:T:G

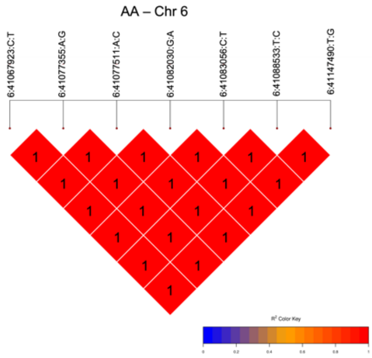
AA – Chr 6
6:41067923:C:T
6:41077355:A:G
6:41077511:A:C
6:41082030:G:A
6:41083056:C:T
6:41088533:T:C
6:41147490:T:G

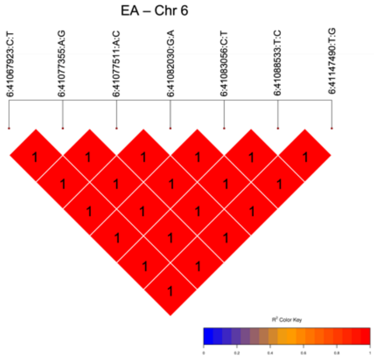
EA – Chr 6
6:41067923:C:T
6:41077355:A:G
6:41077511:A:C
6:41082030:G:A
6:41083056:C:T
6:41088533:T:C
6:41147490:T:G

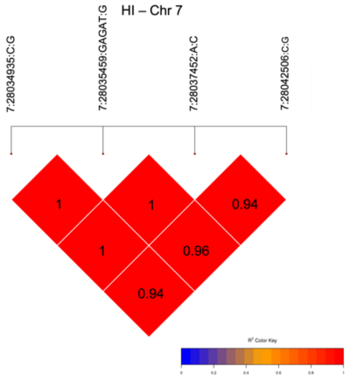
HI – Chr 7
7:28034935:C:G
7:28035459:GAGAT:G
7:28037452:A:C
7:28042506:C:G
1
1
0.94
1
0.96
0.94

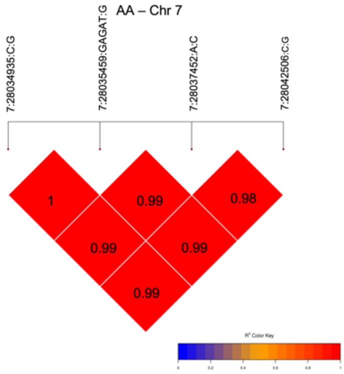
AA – Chr 7
7:28034935:C:G
7:28035459:GAGAT:G
7:28037452:A:C
7:28042506:C:G
1
0.99
0.98
0.99
0.99
0.99

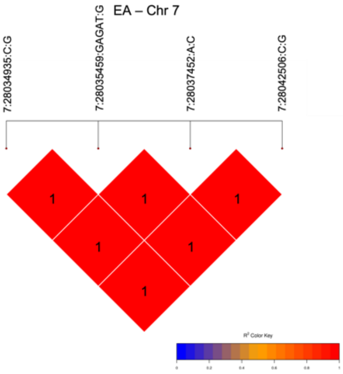
EA – Chr 7
7:28034935:C:G
7:28035459:GAGAT:G
7:28037452:A:C
7:28042506:C:G

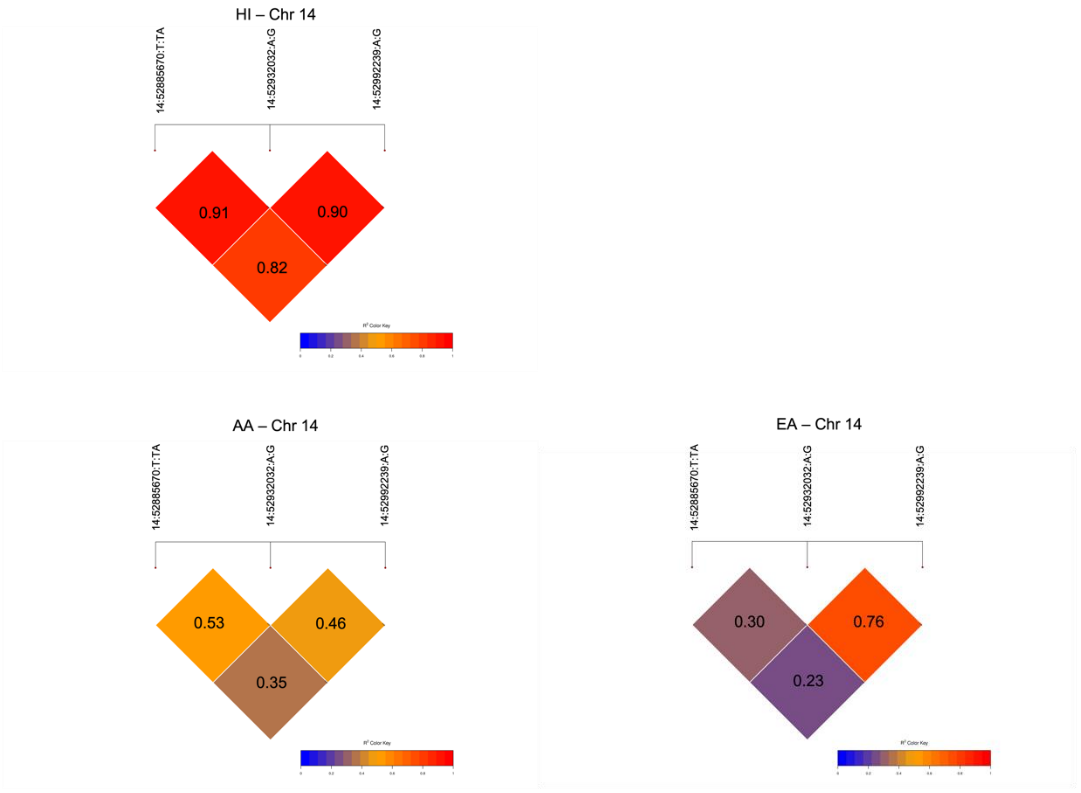


**Figure A3**. LD plots for each of the main GWAS regions with several signals detected with the pooled analysis. LD was calculated using the ADSP data in each population group separately.

EA: White or European ancestry, AA: Black or African-American, HI: Hispanic/Latino

LD: Linkage Disequilibrium

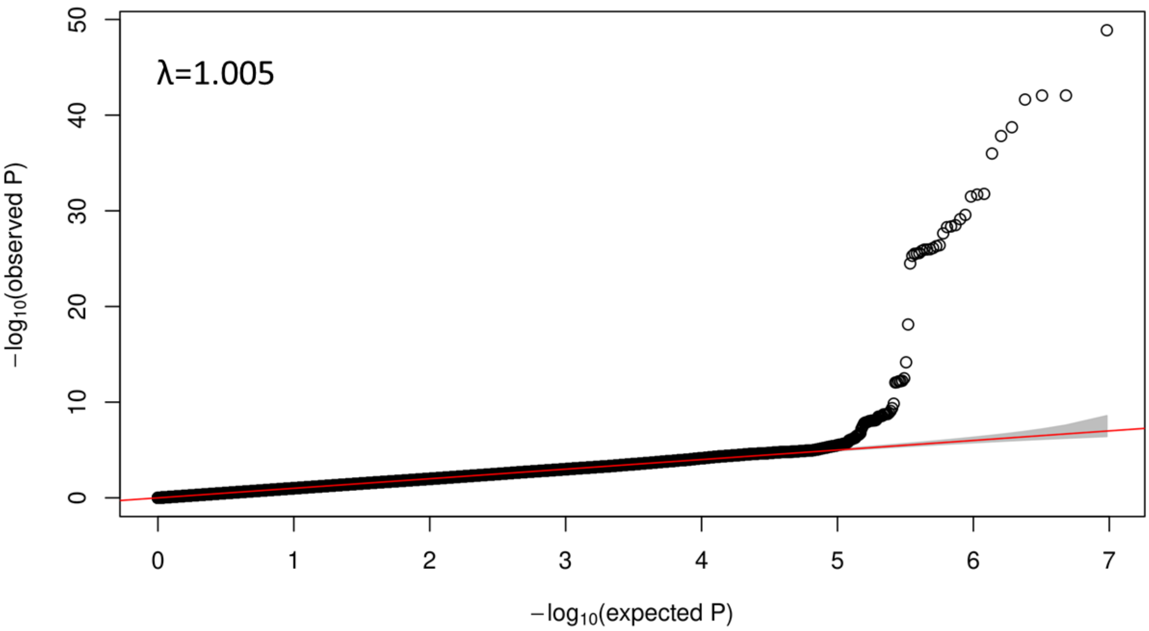


**Figure A4**. QQ-plot of the single-variant association analysis performed in the White/European ancestry group

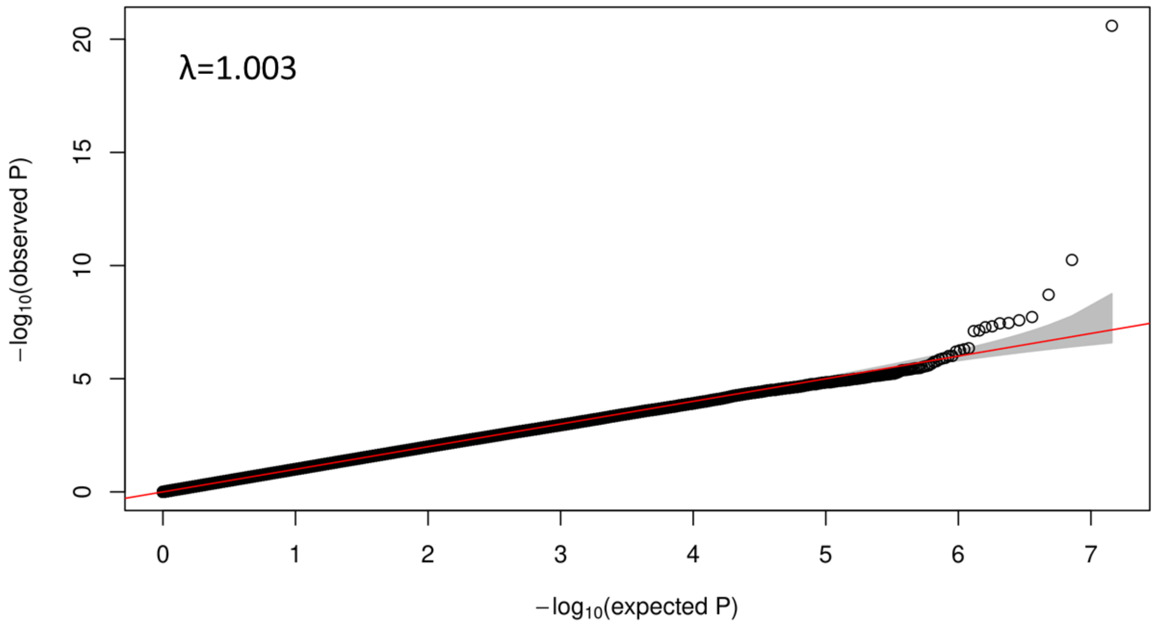


**Figure A5**. QQ-plot of the single-variant association analysis performed in the Black/African-American group

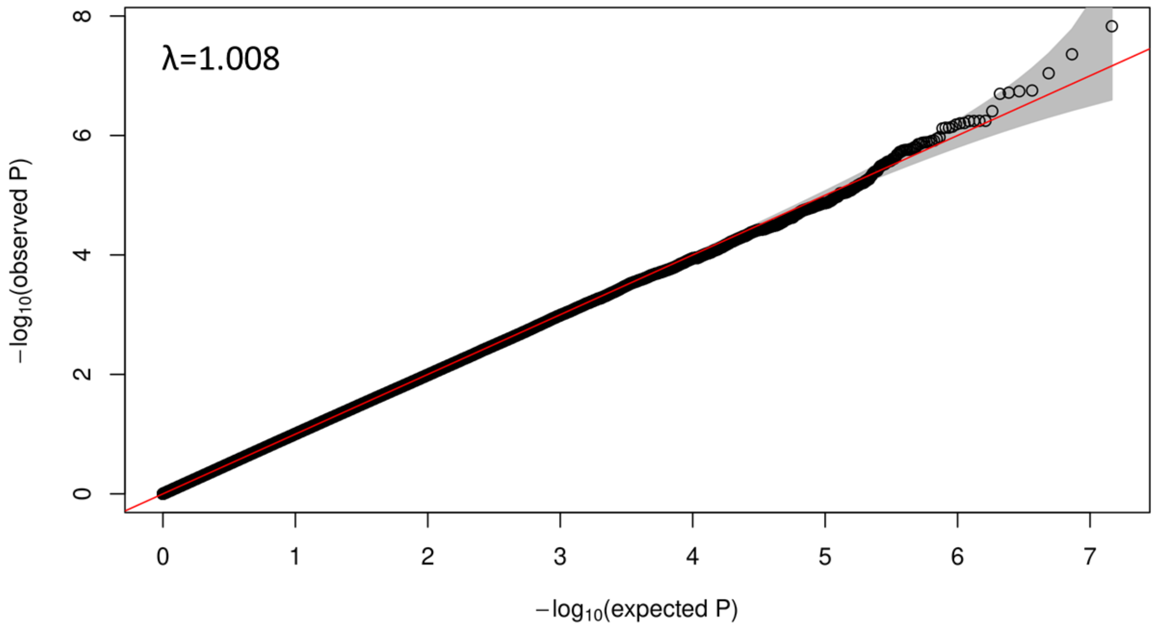


**Figure A6**. QQ-plot of the single-variant association analysis performed in the Hispanic/Latino group

**Fixed Effects model (FE)**

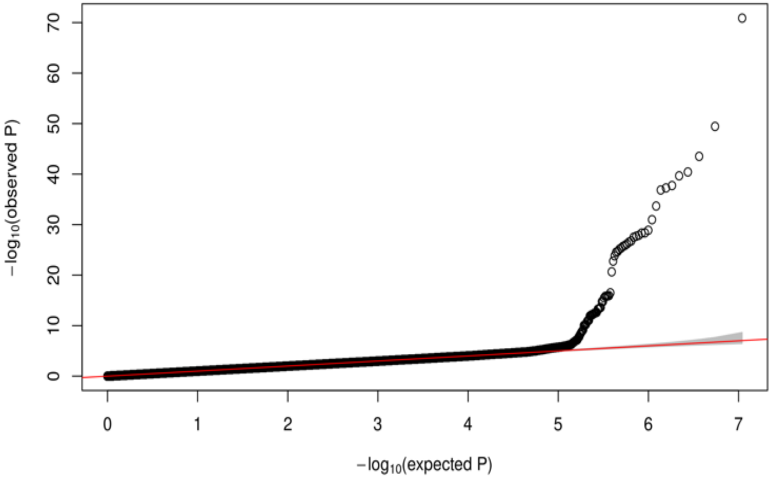


**Random Effects model (RE)**

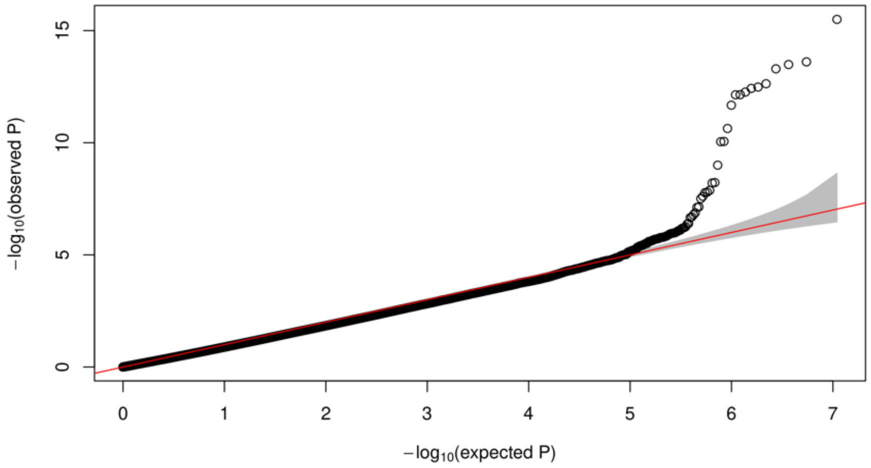


**Han and Eskin's Random Effects model (RE2)**

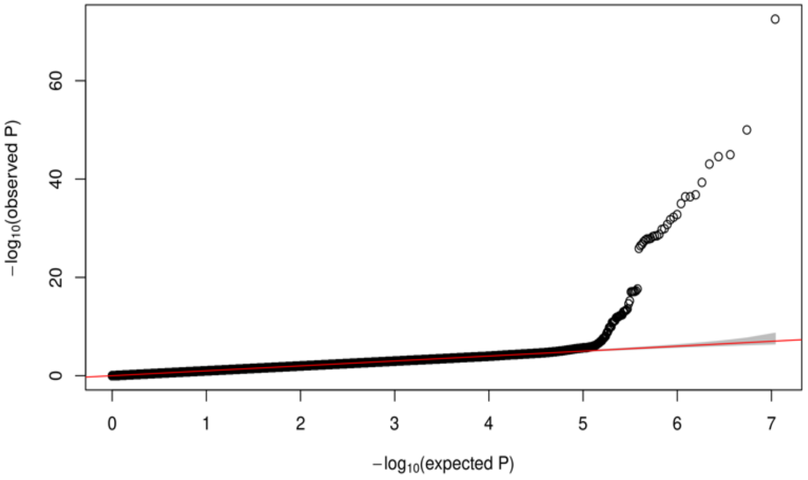


**Figure A7.** QQ-plots for the multi-population meta-analysis under different models.

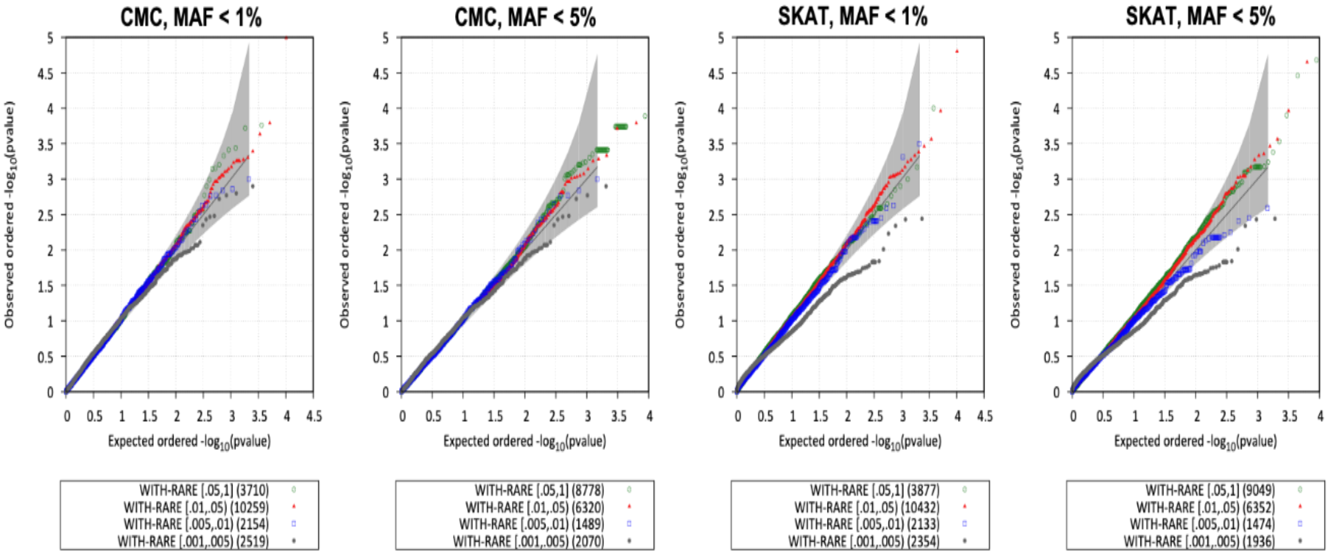


**Figure A8**. QQ plots for the rare-variant aggregation analyses (gene-based) conducted with EPACTS based on missense and loss of function rare genetic variants.

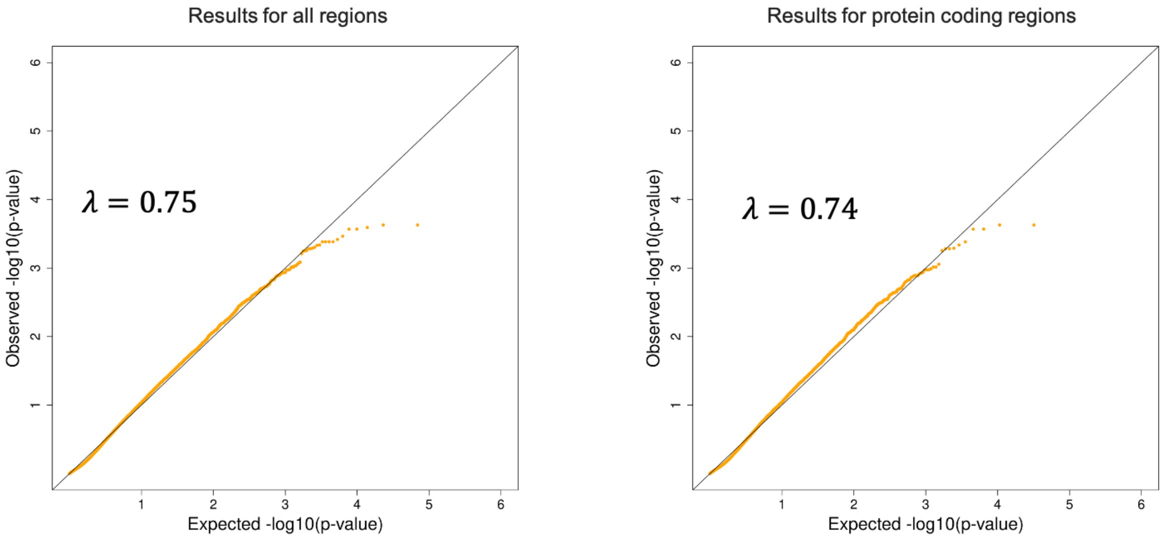


**Figure A9**. QQ plots for the non-coding rare-variant aggregation analysis using STAAR.
